# Knowledge, attitude, and practice regarding COVID-19 outbreak in Bangladesh: An online-based cross-sectional study

**DOI:** 10.1101/2020.05.26.20105700

**Authors:** Most. Zannatul Ferdous, Md. Saiful Islam, Md. Tajuddin Sikder, Abu Syed Md. Mosaddek, J. A. Zegarra-Valdivia, David Gozal

**Author notes:** **Corresponding Author** (DG).

## Abstract

In Bangladesh, an array of measures have been adopted to control the rapid spread of the COVID-19 epidemic. Such general population control measures could significantly influence perception, knowledge, attitudes, and practices (KAP) towards COVID-19. Here, we assessed KAP towards COVID-19 immediately after the lock-down measures were implemented and during the rapid rise period of the outbreak. Online-based cross-sectional study conducted from March 29 to April 19, 2020, involving Bangladeshi residents aged 12–64 years, recruited via social media. After consenting, participants completed an online survey assessing sociodemographic variables, perception, and KAP towards COVID-19. Of the 2017 survey participants, 59.8% were male, the majority were students (71.2%), aged 21-30 years (57.9%), having a bachelor’s degree (61.0%), having family income >30,000 BDT (50.0%), and living in urban areas (69.8). The survey revealed that 48.3% of participants had more accurate knowledge, 62.3% had more positive attitudes, and 55.1% had more frequent practices regarding COVID-19 prevention. Majority (96.7%) of the participants agreed ‘COVID-19 is a dangerous disease’, almost all (98.7%) participants wore a face mask in crowded places, 98.8% agreed to report a suspected case to health authorities, and 93.8% implemented washing hands with soap and water. In multiple logistic regression analyses, COVID-19 more accurate knowledge was associated with age and residence. Sociodemographic factors such as being older, higher education, employment, monthly family income >30,000 BDT, and having more frequent prevention practices were the more positive attitude factors. More frequent prevention practice factors were associated with female sex, older age, higher education, family income > 30,000 BDT, urban area residence, and having more positive attitudes. To improve KAP of general populations is crucial during the rapid rise period of a pandemic outbreak such as COVID-19. Therefore, development of effective health education programs that incorporate considerations of KAP-modifying factors is needed.

## Introduction

Coronavirus disease 2019 (COVID -19) is a global public health threat and has evolved to become a pandemic crisis around the world, which is caused by the severe acute respiratory syndrome, coronavirus 2 (SARS-CoV-2) [1]. In response to this serious situation, COVID-19 was declared as a public health emergency of international concern by the World Health Organization (WHO) on January 30 and called for collaborative efforts of all countries to prevent the rapid spread of COVID-19 [2].In Bangladesh, the first confirmed case was reported on 8 March 2020 [3]. Infection rates apparently remained low until the end of March, but a steep rise in cases began in April 2020 with case doubling times of 2 days [4]. As of 01 June 2020, according to the Institute of Epidemiology, Disease Control and Research (IEDCR), in Bangladesh 49,534 confirmed cases were reported, including 10,597 (21.4%) who recovered, and 672 (1.36%) related deaths [3]. The highest attack rate (AR) was observed in Dhaka city (874.9/1,000,000), followed by (2,040/1,000,000), followed by Narayanganj district (616.2/1,000,000), Munshiganj (432.4/1,000,000), Gazipur (168.7/1,000,000), Gopalganj (145.7/1,000,000), [3].

COVID-19 prompted implementation of public health protocols to control the spread of the virus, many of them involving social distancing, hand washing, and lockdown procedures, but has also resulted in creating public anguish and massive fear [5], particularly among the unaffected population [6]. Bangladesh has not previously experienced epidemics such as SARS or MERS, and it is clear that the public healthcare systems are not readily prepared for COVID-19. The magnitude and rapid proliferation of COVID-19 through slightly symptomatic or asymptomatic infected people in Bangladesh stresses the need to identify the behavioral responses of the population, such as to better address behavioral determinants of pandemic control [7]. Official measures such as school closures, shutdown of offices for an initial 30-day duration, restrictions on leaving home after 6.00 pm, and legal actions on individuals leaving their dwellings after 7.00 pm, along with gathering restrictions in mosques and people gatherings have rapidly been imposed in many regions of the country [8,9]. However, for such measures to be effective, public adherence is essential, which is affected by their knowledge, attitudes, and practices (KAP) towards COVID-19 [10,11]. There are a limited number of studies on knowledge and attitudes during epidemics that have been conducted in Bangladesh. However, the lessons learned from the studies conducted in other countries in an epidemic situation such as the SARS outbreak in 2003 suggest that knowledge and attitudes towards infectious diseases are associated with serious panic and other emotional reactions among the population, which can further complicate attempts to prevent the spread of the disease [12, 13]. Suggestions from a Latin America-based study during the outbreaks of chikungunya, zika, and dengue reported low levels of participation and commitment to the imposed control measures in populations [14].

KAP is an important cognitive key in public health regarding health prevention and promotion. It involves a range of beliefs about the causes of the disease and exacerbating factors, identification of symptoms, and available methods of treatments and consequences [15]. Beliefs about COVID-19 come from different sources, such as stereotypes concerning similar viral diseases, governmental information, social media and internet, previous personal experiences, and medical sources. The accuracy of these beliefs may determine different behaviors about prevention and could vary in the population. In many cases, the absence of knowledge, or if most of these medical-related beliefs are actually false may carry a potential risk factor [16]. In Hubei, China, one of the first studies analyzing attitudes and knowledge about COVID-19 concluded that attitudes towards government measures to contain the epidemic were highly associated with the level of knowledge about COVID-19 [17]. The authors reported that higher levels of information and education were associated with more positive attitudes towards COVID-19 preventive practices [5,17]. Perception of risk is also a key factor in commitment to prevention during outbreaks of global epidemics [5, 18-21].

Considering the lack of studies related to coronavirus epidemics and how to facilitate outbreak management of COVID-19 in Bangladesh, there is an urgent need to understand the public’s KAP of COVID-19. Here, we aimed to investigate KAP towards COVID-19 during the rapid rise period and immediately after the implementation of lockdown measures in Bangladesh.

## Methods

### Participation and procedure

A cross-sectional and anonymous online population-based survey was conducted among individuals aged 12–64 years. The survey was conducted from March 29 to April 20, 2020, immediately after the implementation of lockdown measures by the government of Bangladesh. A semi-structured questionnaire was designed for the Google survey tool (Google Forms), and the generated link was shared to public on social media (i.e., Facebook, WhatsApp). The link was also shared personally to the contact list of investigators and research assistants. The investigators’ decision to collect the data using online approaches was predicated on maintaining social distance during the strict lockdown in Bangladesh. Initially, 2,068 potential respondents provided informed consent. Of these, 2,017 respondents completed the entire survey, generating a response rate of 97.5%. The inclusion criteria to participate in the study were being a Bangladeshi resident, having internet access, and volunteering.

### Measures

A semi-structured and self-reported questionnaire containing informed consent, questions regarding socio-demographics, knowledge, attitude, and practice.

#### Socio-demographic measures

Socio-demographic information was collected, including gender, age, education, occupation, marital status, nature of the family (nuclear/joint, with the joint being an extended family, often of multiple generations), number of family members, monthly family income, and location of permanent residence.

#### Knowledge, attitude, and practice

To assess the level of knowledge, attitude, and practice of the respondents, a total of 17 questions (including 6 for knowledge, 6 for attitude, and 7 for practice) were included. The survey questions were adapted and modified from previously published literature regarding viral epidemics related to MERS-CoV disease [23,24], infection prevention and control measures for COVID-19 by World Health Organization [25], and guidelines suggested by the country’s Institute of Epidemiology, Disease Control and Research (IEDCR) [26].

After completion of the initial draft of the survey questionnaire, it was validated and adopted as follows: Firstly, the questionnaire was sent to four experts. After coordination and consensus of all experts’ opinions, the final questionnaire was drafted, and underwent pilot testing in 30 individuals to confirm the reliability of the questionnaire. The data from the pilot study were loaded into SPSS version 25, and subjected to reliability coefficient analysis. The Cronbach’s alpha coefficient of the KAP questions was 0.73, which indicates acceptable internal consistency [27].

The knowledge section consisted of 6 items of a and each question was responded as *“Yes’”, “No”* and “*Don’t know*”. The correct answer was coded as 1, while the wrong answer was coded as 0. The total score ranged from 0-6, with an overall greater score indicates more accurate knowledge. A cut off level of ≥4 was set for more accurate knowledge.

The attitude section consisted of 6 items, and the response of each item was responded on a 3-point Likert scale as follows 0 (“*Disagree*”), 1 (“*Undecided*”), and *2(“Agree”)*. The total score ranges from 0 to 12, with an overall greater score indicates more positive attitudes towards the COVID-19. A cut off level of ≥11was set for more positive attitudes towards the prevention of COVID-19.

The practice section included 7 items practice measures responding to the COVID-19, and each item was responded as “*Yes*”, “*No*”, and “*Sometimes*”. Practice items’ total score ranges from 0-7, with an overall greater score indicates more frequent practices towards the COVID-19. A cut off level of ≥6was set for more frequent practices.

### Statistical analysis

The data analysis was performed using Microsoft Excel 2019 and SPSS version 25.0 (Chicago, IL, USA). Microsoft Excel was used for editing, sorting, and coding. The excel file was then imported into SPSS software. Descriptive statistics (frequencies, percentages, means, standard deviations) and first-order analyses (i.e., chi-square tests) were performed. Binary logistic regression was performed with a 95% confidence interval to determine significant associations between categorical dependent and independent variables.

### Ethical considerations

The study was conducted in accordance with the Institutional Research Ethics and the declaration of Helsinki. Formal ethical approval was granted by the Ethical Review Committee, Uttara Adhunik Medical College, Uttara, Dhaka-1260, Bangladesh (Ref: UAMC/ERC/04/2020). The consent form documented the aims, nature, and procedure of the study. Anonymity and confidentially were strictly maintained.

## Results

A total of 2017 respondents were included in the final analysis, of which 59.8% male with an average age was 24.4±5.4 years (SD) ranging from 12 to 64 years. Almost all respondents were not married (80.8%). The majority were students (71.2%), had a bachelor’s level of education (61.0%), came from urban areas (69.8%), lived in nuclear families (77.9%) and their monthly family income was >30,000 BDT (50.0%) (Table 1).

**Table 1.**
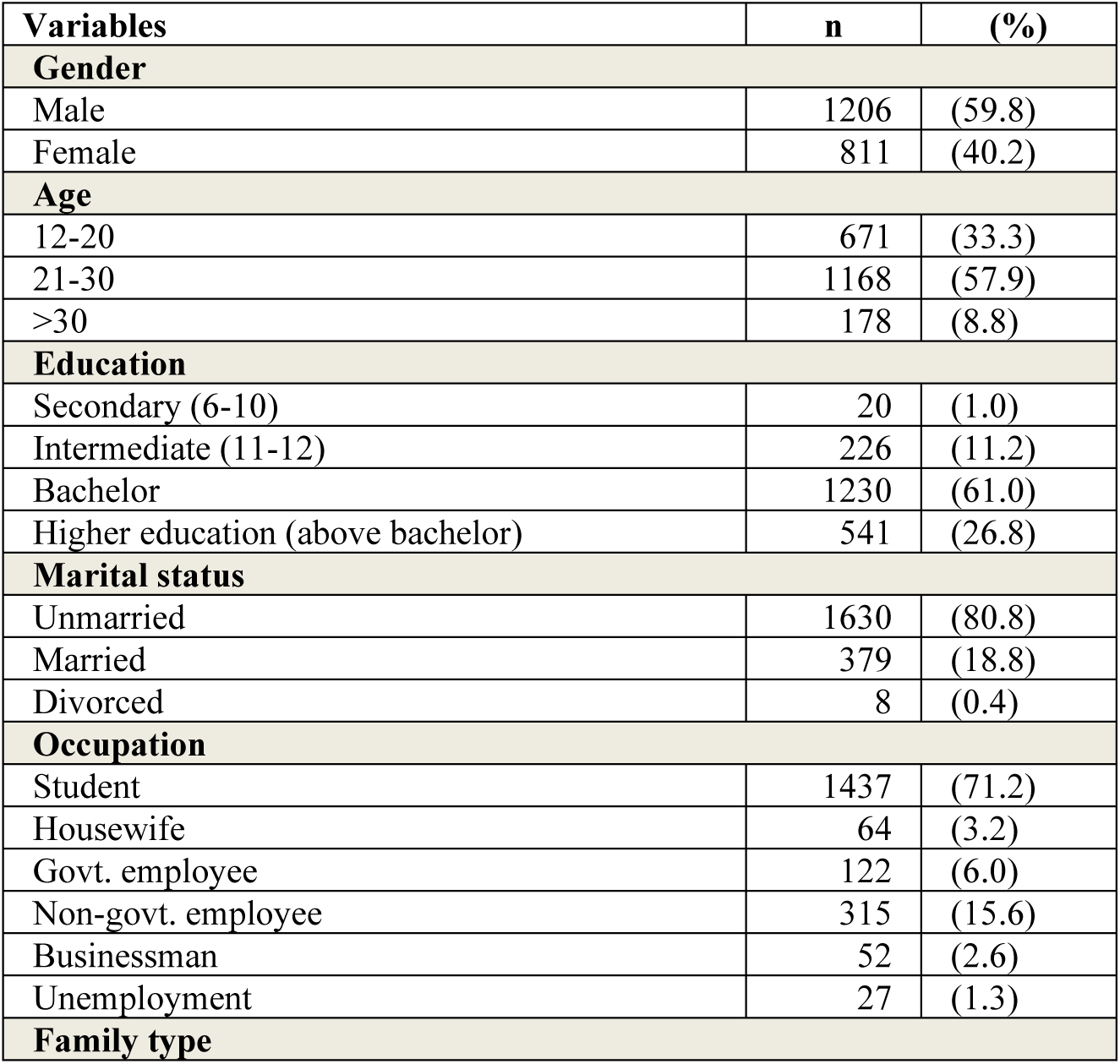
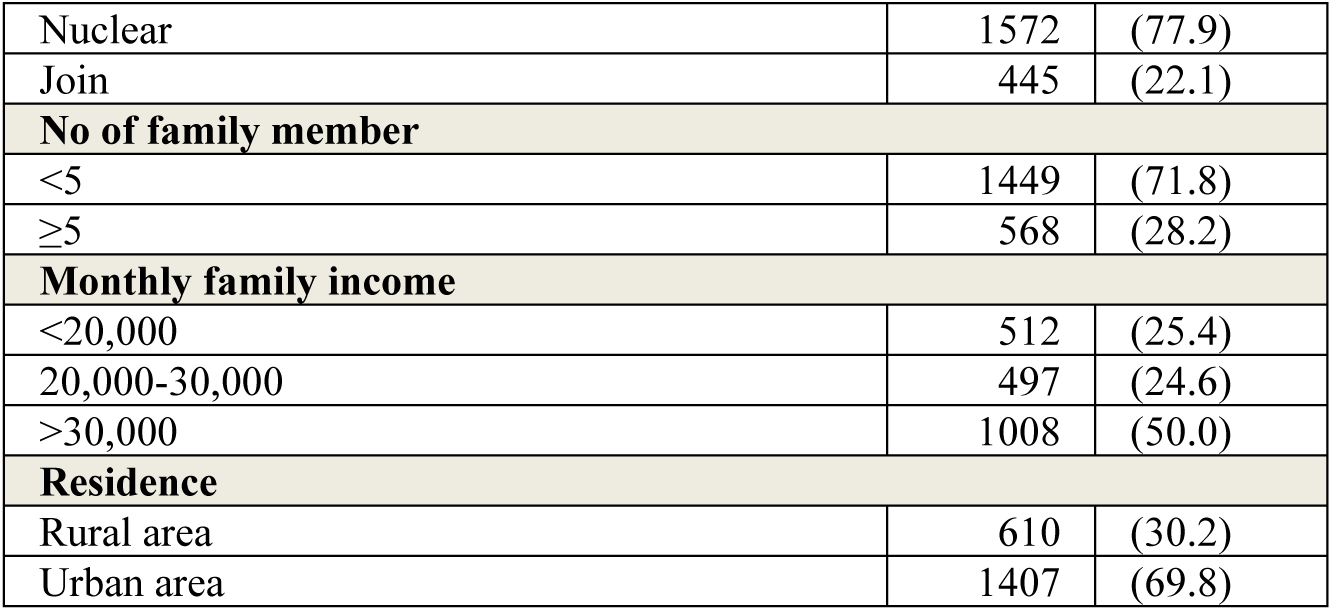
Demographic characteristics of participants (N=2017)

| Variables | n | (%) |
| --- | --- | --- |
| **Gender** |  |  |
| Male | 1206 | (59.8) |
| Female | 811 | (40.2) |
| **Age** |  |  |
| 12-20 | 671 | (33.3) |
| 21-30 | 1168 | (57.9) |
| >30 | 178 | (8.8) |
| **Education** |  |  |
| Secondary (6-10) | 20 | (1.0) |
| Intermediate (11-12) | 226 | (11.2) |
| Bachelor | 1230 | (61.0) |
| Higher education (above bachelor) | 541 | (26.8) |
| **Marital status** |  |  |
| Unmarried | 1630 | (80.8) |
| Married | 379 | (18.8) |
| Divorced | 8 | (0.4) |
| **Occupation** |  |  |
| Student | 1437 | (71.2) |
| Housewife | 64 | (3.2) |
| Govt. employee | 122 | (6.0) |
| Non-govt. employee | 315 | (15.6) |
| Businessman | 52 | (2.6) |
| Unemployment | 27 | (1.3) |
| **Family type** |  |  |
| Nuclear | 1572 | (77.9) |
| Join | 445 | (22.1) |
| **No of family member** |  |  |
| <5 | 1449 | (71.8) |
| ≥5 | 568 | (28.2) |
| **Monthly family income** |  |  |
| <20,000 | 512 | (25.4) |
| 20,000-30,000 | 497 | (24.6) |
| >30,000 | 1008 | (50.0) |
| **Residence** |  |  |
| Rural area | 610 | (30.2) |
| Urban area | 1407 | (69.8) |

### Perception towards the COVID-19 about mode of transmission, incubation period, symptoms, risk factors, treatments, prevention, initiatives, and challenges

In the perception component, Table 2 depicts our findings. For the mode of transmission, more than half of the respondents reported close contact with an infected person (93.7%), direct transmission during coughing (66.4%), touching contaminated surfaces (61.3%), along with others as just as contact with infected animals (30.8%), through eating infected animal products (e.g., meat, milk) (21.4%), and only 0.5% had no idea about the mode of transmission of COVID-19. Most of the respondents (91.3%) reported the correct incubation period (2-14 days), and only 2.4% had no knowledge. Most of the respondents (99.4%) reported fever, dry cough, and difficulty breathing as the common symptoms of the COVID-19. On the other hand, half of the respondents (51.2%) reported sore throat, nasal stuffiness, along with headache (0.1%), diarrhea (0.7%), and no idea (0.4%).

**Table 2.**
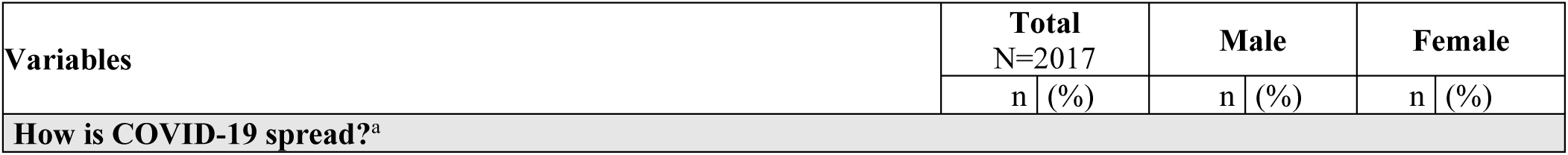
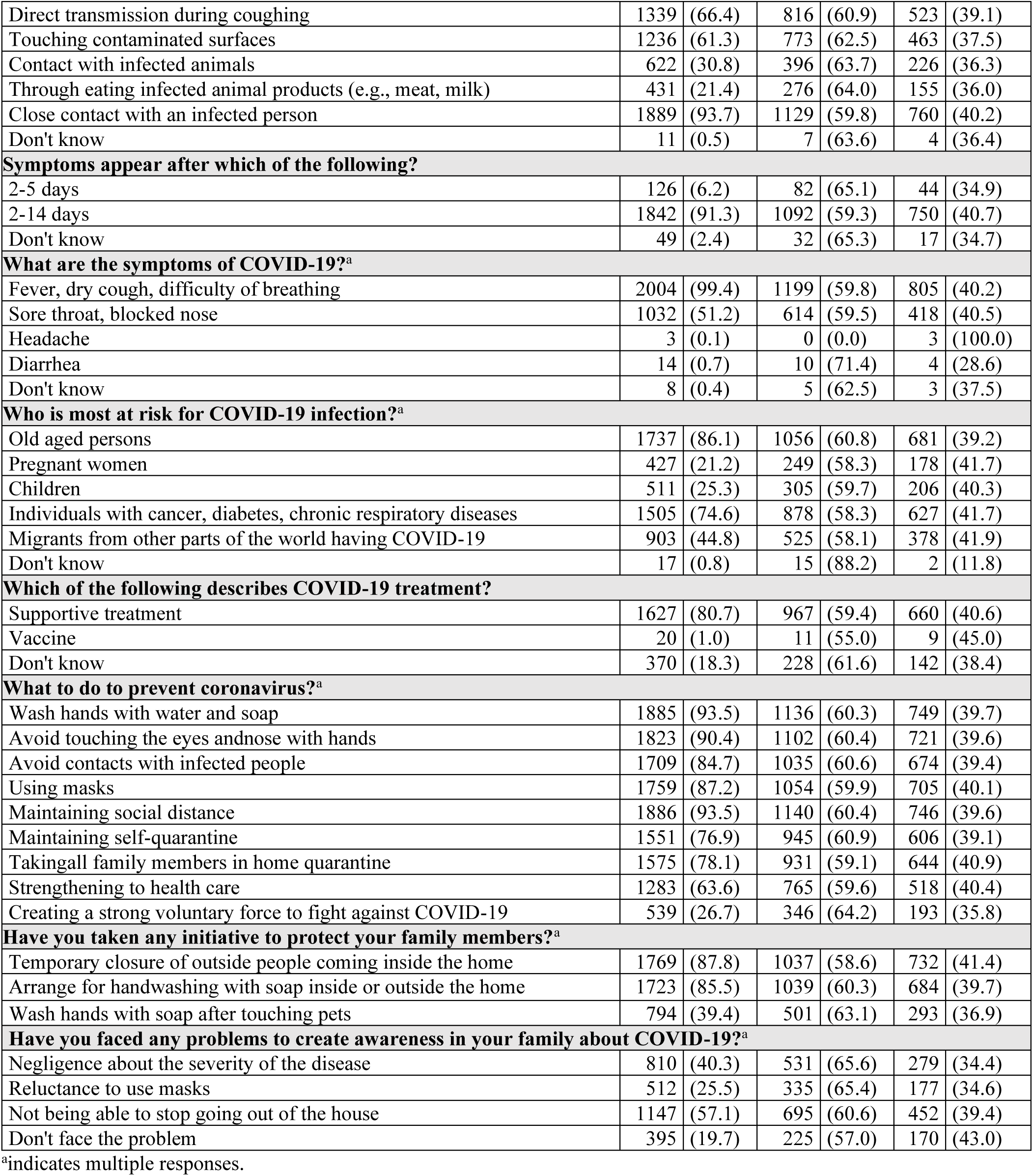
Perception towards COVID-19 about the mode of transmission, incubation period, symptoms, risk factors, treatments, prevention initiatives, and challenges.

The respondents identified risk groups for developing COVID-19 as follows: older age persons (86.1%), individuals with cancer, diabetes, chronic respiratory diseases (74.6%), migrants from other parts of the world having COVID-19 (44.8%), children (25.3%), pregnant women (21.2%), and no idea (0.8%). The majority (80.7%) reported supportive treatments, but a vaccine was rarely mentioned (1.0%), and 18.3% had no idea about the treatment options of COVID-19.

The respondents recognized the following preventive measures for the COVID-19: washing hands with water and soap (93.5%), maintaining social distance (93.5%), avoid touching the eyes, nose with hands (90.4%), using a mask (87.2%), avoid contacts with infected people (84.7%), taking all family members into home quarantine (78.1%), maintaining self-quarantine (76.9%), strengthening to health care (63.6%), and creating a strong force to fight against COVID-19 (26.7%).

The respondents took the initiative to protect their family members: temporary and absolute restricted access to outside people coming inside the home (87.8%), arrange for handwashing with soap inside or outside the home (85.5%), and wash hands with soap after touching pets (39.4%). The respondents also reported that they faced many problems to create awareness among their family members: not being able to stop from leaving the house (57.1%), negligence about the severity of the disease (40.3%), reluctance to use masks (25.5%), and only 19.7% had no problems.

### Knowledge

For each question of knowledge, the distribution of responses from participants is presented in Table 3 with gender differences. There were no significant gender differences for each item of knowledge questions; 48.3% of respondents had more accurate knowledge, and 51.7% of respondents had comparatively inaccurate knowledge regarding COVID-19. The rates of adequate knowledge were significantly more likely to be among (i)younger (12-20 years) (49.3% vs. 38.8% in aged more than 30 years, *p*=.029), and (ii) be a respondent from a rural area (52.8% vs. 46.3% in those from an urban area, *p*=.008).

**Table 3.** Knowledge and gender difference of participants (N=2017)

| Variables | Total N=2017 | | Male | | Female | | $\chi^2$ | df | p-value |
| --- | --- | --- | --- | --- | --- | --- | --- | --- | --- |
|  | n | (%) | n | (%) | n | (%) |  |  |  |
| Is COVID-19 a dangerous disease? |  |  |  |  |  |  |  |  |  |
| Yes | 1951 | (96.7) | 1160 | (96.2) | 791 | (97.5) | 2.988 | 2 | 0.224 |
| No | 40 | (2.0) | 27 | (2.2) | 13 | (1.6) |  |  |  |
| Don't know | 26 | (1.3) | 19 | (1.6) | 7 | (0.9) |  |  |  |
| Does it affect only humans? |  |  |  |  |  |  |  |  |  |
| Yes | 1210 | (60.0) | 735 | (60.9) | 475 | (58.6) | 1.161 | 2 | 0.560 |
| No | 567 | (28.1) | 330 | (27.4) | 237 | (29.2) |  |  |  |
| Don't know | 240 | (11.9) | 141 | (11.7) | 99 | (12.2) |  |  |  |
| Does it transmit from humans to animals? |  |  |  |  |  |  |  |  |  |
| Yes | 1013 | (50.2) | 612 | (50.7) | 401 | (49.4) | 4.690 | 2 | 0.096 |
| No | 578 | (28.7) | 358 | (29.7) | 220 | (27.1) |  |  |  |
| Don't know | 426 | (21.1) | 236 | (19.6) | 190 | (23.4) |  |  |  |
| Does it transmit from animals to humans? |  |  |  |  |  |  |  |  |  |
| Yes | 1013 | (50.2) | 612 | (50.7) | 401 | (49.4) | 4.690 | 2 | 0.096 |
| No | 578 | (28.7) | 358 | (29.7) | 220 | (27.1) |  |  |  |
| Don't know | 426 | (21.1) | 236 | (19.6) | 190 | (23.4) |  |  |  |
| Is it transmitted by animal products (e.g., milk, meat)? |  |  |  |  |  |  |  |  |  |
| Yes | 509 | (25.2) | 300 | (24.9) | 209 | (25.8) | 0.406 | 2 | 0.816 |
| No | 1017 | (50.4) | 615 | (51.0) | 402 | (49.6) |  |  |  |
| Don't know | 491 | (24.3) | 291 | (24.1) | 200 | (24.7) |  |  |  |
| Is it transmitted in well-cooked products? |  |  |  |  |  |  |  |  |  |
| Yes | 53 | (2.6) | 35 | (2.9) | 18 | (2.2) | 3.859 | 2 | 0.145 |
| No | 1821 | (90.3) | 1076 | (89.2) | 745 | (91.9) |  |  |  |
| Don't know | 143 | (7.1) | 95 | (7.9) | 48 | (5.9) |  |  |  |

The sociodemographic factors of more accurate knowledge were 12-29 years age group vs. >30 years (OR=1.54; 95%CI=1.10-2.16, *p*=.012), and rural vs. urban areas (OR=1.295; 95CI%=1.07-1.57, *p*=008).

### Attitude

For each question focused on attitude, the distribution of responses from participants is presented in Table 4. The response rates of “*Agree*” were significantly higher in females (99.5% vs. 98.3% in males, *p*=.043) to the item of attitude section regarding *“It is crucial to report a suspected case to health authorities’”*. Furthermore, the response rates of “*Agree*” were significantly higher in females (99.6% vs. 98.1% in males, *p*=.011) to the item of attitude section regarding *“It is important to use a face mask in a crowded place.”*

**Table 4.** Attitude and gender difference of participants (N=2017)

| Variables | Total N=2017 | | Male | | Female | | $\chi^2$ | df | p-value |
| --- | --- | --- | --- | --- | --- | --- | --- | --- | --- |
|  | n | (%) | n | (%) | n | (%) |  |  |  |
| It is crucial to report a suspected case to health authorities. |  |  |  |  |  |  |  |  |  |
| Agree | 1993 | (98.8) | 1186 | (98.3) | 807 | (99.5) | 6.292 | 2 | 0.043 |
| Undecided | 12 | (0.6) | 11 | (0.9) | 1 | (0.1) |  |  |  |
| Disagree | 12 | (0.6) | 9 | (0.7) | 3 | (0.4) |  |  |  |
| It is important to use a face mask in crowded place. |  |  |  |  |  |  |  |  |  |
| Agree | 1991 | (98.7) | 1183 | (98.1) | 808 | (99.6) | 9.053 | 2 | 0.011 |
| Undecided | 11 | (0.5) | 10 | (0.8) | 1 | (0.1) |  |  |  |
| Disagree | 15 | (0.7) | 13 | (1.1) | 2 | (0.2) |  |  |  |
| It is important to wash hands and face after coming outsides. |  |  |  |  |  |  |  |  |  |
| Agree | 2006 | (99.5) | 1197 | (99.3) | 809 | (99.8) | 3.492 | 2 | 0.174 |
| Undecided | 5 | (0.2) | 5 | (0.4) | 0 | (0.0) |  |  |  |
| Disagree | 6 | (0.3) | 4 | (0.3) | 2 | (0.2) |  |  |  |
| COVID-19 is a preventable disease. |  |  |  |  |  |  |  |  |  |
| Agree | 1829 | (90.7) | 1091 | (90.5) | 738 | (91.0) | 4.211 | 2 | 0.122 |
| Undecided | 109 | (5.4) | 60 | (5.0) | 49 | (6.0) |  |  |  |
| Disagree | 79 | (3.9) | 55 | (4.6) | 24 | (3.0) |  |  |  |
| It can be treated at home. |  |  |  |  |  |  |  |  |  |
| Agree | 1158 | (57.4) | 686 | (56.9) | 472 | (58.2) | 1.995 | 2 | 0.369 |
| Undecided | 240 | (11.9) | 137 | (11.4) | 103 | (12.7) |  |  |  |
| Disagree | 619 | (30.7) | 383 | (31.8) | 236 | (29.1) |  |  |  |
| Health education can play an important role in COVID-19 prevention. |  |  |  |  |  |  |  |  |  |
| Agree | 1928 | (95.6) | 1148 | (95.2) | 780 | (96.2) | 1.805 | 2 | 0.406 |
| Undecided | 52 | (2.6) | 32 | (2.7) | 20 | (2.5) |  |  |  |
| Disagree | 37 | (1.8) | 26 | (2.2) | 11 | (1.4) |  |  |  |

The findings indicated that 62.3% of respondents had more positive attitudes towards COVID-19. The rates of more positive attitudes were significantly more likely to be (i) among older individuals (> 30 years) (72.5% vs. 55.1% in aged 12-20 years, *p*<.001), and (ii) those with higher education (74.1% vs. 52.2% in intermediate [class 11-12], *p*<.001), (iii)mamed (70.4% vs. 37.5% in divorced, *p*=.001), (iv)housewives (78.1% vs. 58.2% in student, *p*<.001), (v) come from joint family (66.7% vs. 61.1% in nuclear family, *p*=.029), (vi) have monthly family income > 30,000 BDT (65.2% vs. 57.8% in those less than 20,000 BDT, *p*=.016), and (vii)have more frequent safety-related preventive practices(66.1% vs. 57.7% in comparatively less frequent practices, *p*<.001).

Finally, regarding variables related to more positive attitudes against COVID-19, we found being younger (aged 12-20 years) vs. older (>30 years) (OR=0.47; 95%CI=0.33-.67, *p*<.001). Additional factors of more positive attitudes against COVID-19 were having higher education (above bachelor), being unemployed, having joint families, having monthly family income more than 30,000 BDT, and having more frequent practices (Table 6).

### Practice

For each question-related to practice, the distribution of responses from participants is presented in Table 5. The response rates of “*Yes*” were significantly higher in females (81.8% vs. 73.5% in males, *p*<.001) to the item of practice section regarding “*Do you use tissues during coughing/sneezing*?”, as well as “*Do you wash hands frequently using water and soaps?”* (95.6% vs. 92.5% in males, *p*=.023). Similarly, “*Yes*” response rates were significantly higher in females (96.2% vs. 87.1% in males, *p*<.001) to “*Do you maintain social distance (or home quarantine)?*”, to *“Do you maintain a healthy lifestyle focusing on outbreak?”*(88.3% vs. 81.4% in males, *p*<.001), and to “*Do you obey all government rules related to the COVID-19?”* (93.0% vs. 85.4% in males, *p*<.001).

**Table 5.** Practice and gender difference of participants (N=2017)

| Variables | Total N=2017 | | Male | | Female | | $\chi^2$ | df | p-value |
| --- | --- | --- | --- | --- | --- | --- | --- | --- | --- |
|  | n | (%) | n | (%) | n | (%) |  |  |  |
| Do you use tissues or hanker chips during coughing/sneezing? |  |  |  |  |  |  |  |  |  |
| Yes | 1549 | (76.8) | 886 | (73.5) | 663 | (81.8) | 23.392 | 2 | <0.001 |
| No | 74 | (3.7) | 59 | (4.9) | 15 | (1.8) |  |  |  |
| Sometimes | 394 | (19.5) | 261 | (21.6) | 133 | (16.4) |  |  |  |
| Do you wash hands frequently using water and soaps? |  |  |  |  |  |  |  |  |  |
| Yes | 1891 | (93.8) | 1116 | (92.5) | 775 | (95.6) | 7.570 | 2 | 0.023 |
| No | 14 | (0.7) | 10 | (0.8) | 4 | (0.5) |  |  |  |
| Sometimes | 112 | (5.6) | 80 | (6.6) | 32 | (3.9) |  |  |  |
| Do you avoid touching face and eyes? |  |  |  |  |  |  |  |  |  |
| Yes | 1228 | (60.9) | 734 | (60.9) | 494 | (60.9) | 0.118 | 2 | 0.943 |
| No | 154 | (7.6) | 94 | (7.8) | 60 | (7.4) |  |  |  |
| Sometimes | 635 | (31.5) | 378 | (31.3) | 257 | (31.7) |  |  |  |
| Do you maintain social distance (or home quarantine)? |  |  |  |  |  |  |  |  |  |
| Yes | 1831 | (90.8) | 1051 | (87.1) | 780 | (96.2) | 47.237 | 2 | <0.001 |
| No | 41 | (2.0) | 34 | (2.8) | 7 | (0.9) |  |  |  |
| Sometimes | 145 | (7.2) | 121 | (10.0) | 24 | (3.0) |  |  |  |
| Do you eat healthy food focusing on outbreak? |  |  |  |  |  |  |  |  |  |
| Yes | 760 | (37.7) | 464 | (38.5) | 296 | (36.5) | 0.806 | 1 | 0.369 |
| No | 1257 | (62.3) | 742 | (61.5) | 515 | (63.5) |  |  |  |
| Sometimes | 0 | (0.0) | 0 | (0.0) | 0 | (0.0) |  |  |  |
| **Do you maintain a healthy lifestyle focusing on outbreak?** |  |  |  |  |  |  |  |  |  |
| Yes | 1698 | (84.2) | 982 | (81.4) | 716 | (88.3) | 17.779 | 2 | <0.001 |
| No | 19 | (0.9) | 15 | (1.2) | 4 | (0.5) |  |  |  |
| Sometimes | 300 | (14.9) | 209 | (17.3) | 91 | (11.2) |  |  |  |
| **Do you obey all government rules related to the COVID?** |  |  |  |  |  |  |  |  |  |
| Yes | 1784 | (88.4) | 1030 | (85.4) | 754 | (93.0) | 29.710 | 2 | <0.001 |
| No | 39 | (1.9) | 25 | (2.1) | 14 | (1.7) |  |  |  |
| Sometimes | 194 | (9.6) | 151 | (12.5) | 43 | (5.3) |  |  |  |

**Table 6.** Distribution and risk factors of Knowledge, Attitude and Practice among participants.

| Variables | Knowledge |  |  |  |  |  |  |  | Attitudes |  |  |  |  |  |  | Practices |  |  |  |  |  |  |  |  |
| --- | --- | --- | --- | --- | --- | --- | --- | --- | --- | --- | --- | --- | --- | --- | --- | --- | --- | --- | --- | --- | --- | --- | --- | --- |
| | Less accurate | | More accurate | | $\chi^2$<br>*p-value* | OR | 95%CI | *p-value* | Less positive | | More positive | | $\chi^2$<br>*p-value* | OR | 95%CI | *p-value* | Less frequent | | More frequent | | $\chi^2$<br>*p-value* | OR | 95%CI | *p-value* |
|  | N | (%) | N | (%) |  |  |  |  | N | (%) | N | (%) |  |  |  |  | N | (%) |  |  |  |  |  |  |
| Gender |  |  |  |  |  |  |  |  |  |  |  |  |  |  |  |  |  |  |  |  |  |  |  |  |
| Male | 626 | (51.9) | 580 | (48.1) | 0.046 | 0.981 | 0.821-1.172 | 0.829 | 472 | (39.1) | 734 | (60.9) | 2.715 | 0.856 | 0.712-1.030 | 0.100 | 572 | (47.4) | 634 | (52.6) | 8.583 | 0.764 | 0.638-0.915 | 0.003 |
| Female | 417 | (51.4) | 394 | (48.6) | 0.829 | 1 |  |  | 288 | (35.5) | 523 | (64.5) | 0.099 | 1 |  |  | 331 | (40.8) | 480 | (59.2) | 0.003 | 1 |  |  |
| Age (years) |  |  |  |  |  |  |  |  |  |  |  |  |  |  |  |  |  |  |  |  |  |  |  |  |
| 12-20 | 340 | (50.7) | 331 | (49.3) | 7.100 | 1.538 | 1.098-2.155 | 0.012 | 301 | (44.9) | 370 | (55.1) | 25.841 | 0.467 | 0.325-.671 | <0.001 | 345 | (51.4) | 326 | (48.6) | 20.474 | 0.530 | 0.377-0.746 | <0.001 |
| 21-30 | 594 | (50.9) | 574 | (49.1) | 0.029 | 1.527 | 1.106-2.108 | 0.010 | 410 | (35.1) | 758 | (64.9) | <0.001 | 0.702 | 0.495-.997 | 0.048 | 494 | (42.3) | 674 | (57.7) | <0.001 | 0.766 | 0.552-1.063 | 0.110 |
| >30 | 109 | (61.2) | 69 | (38.8) |  | 1 |  |  | 49 | (27.5) | 129 | (72.5) |  | 1 |  |  | 64 | (36.0) | 114 | (64.0) |  | 1 |  |  |
| Education |  |  |  |  |  |  |  |  |  |  |  |  |  |  |  |  |  |  |  |  |  |  |  |  |
| Secondary (6-10) | 8 | (40.0) | 12 | (60.0) | 4.347 | 1.419 | 0.571-3.527 | 0.451 | 9 | (45.0) | 11 | (55.0) | 47.791 | 0.427 | 0.173-1.051 | 0.064 | 13 | (65.0) | 7 | (35.0) | 23.415 | 0.308 | 0.121-0.786 | 0.014 |
| Intermediate (11-12) | 117 | (51.8) | 109 | (48.2) | 0.226 | 0.881 | 0.646-1.202 | 0.426 | 108 | (47.8) | 118 | (52.2) | <0.001 | 0.381 | 0.276-0.528 | <0.001 | 110 | (48.7) | 116 | (51.3) | <0.001 | 0.604 | 0.441-0.827 | 0.002 |
| Bachelor | 655 | (53.3) | 575 | (46.7) |  | 0.830 | 0.678-1.017 | 0.072 | 503 | (40.9) | 727 | (59.1) |  | 0.505 | 0.404-0.631 | <0.001 | 583 | (47.4) | 647 | (52.6) |  | 0.636 | 0.516-0.782 | <0.001 |
| Higher education (above bachelor) | 263 | (48.6) | 278 | (51.4) |  | 1 |  |  | 140 | (25.9) | 401 | (74.1) |  | 1 |  |  | 197 | (36.4) | 344 | (63.6) |  | 1 |  |  |
| Marital status |  |  |  |  |  |  |  |  |  |  |  |  |  |  |  |  |  |  |  |  |  |  |  |  |
| Single | 835 | (51.2) | 795 | (48.8) | 1.659 | 0.571 | 0.136-2.398 | 0.444 | 643 | (39.4) | 987 | (60.6) | 14.932 | 2.558 | 0.609-10.742 | 0.199 | 751 | (46.1) | 879 | (53.9) | 5.854 | 0.702 | 0.167-2.948 | 0.629 |
| Married | 205 | (54.1) | 174 | (45.9) | 0.436 | 0.509 | 0.120-2.161 | 0.360 | 112 | (29.6) | 267 | (70.4) | 0.001 | 3.973 | 0.934-16.909 | 0.062 | 149 | (39.3) | 230 | (60.7) | 0.054 | 0.926 | 0.218-3.933 | 0.917 |
| Divorced | 3 | (37.5) | 5 | (62.5) |  | 1 |  |  | 5 | (62.5) | 3 | (37.5) |  | 1 |  |  | 3 | (37.5) | 5 | (62.5) |  | 1 |  |  |
| Occupation |  |  |  |  |  |  |  |  |  |  |  |  |  |  |  |  |  |  |  |  |  |  |  |  |
| Student | 733 | (51.0) | 704 | (49.0) | 5.584 | 1.633 | 0.743-3.590 | 0.223 | 601 | (41.8) | 836 | (58.2) | 40.508 | 0.397 | 0.159-0.991 | 0.048 | 687 | (47.8) | 750 | (52.2) | 20.203 | 0.873 | 0.406-1.879 | 0.729 |
| Housewife | 40 | (62.5) | 24 | (37.5) | 0.349 | 1.020 | 0.402-2.587 | 0.967 | 14 | (21.9) | 50 | (78.1) | <0.001 | 1.020 | 0.345-3.016 | 0.971 | 20 | (31.3) | 44 | (68.8) | 0.001 | 1.760 | 0.698-4.438 | 0.231 |
| Govt. employee | 59 | (48.4) | 63 | (51.6) |  | 1.815 | 0.770-4.281 | 0.173 | 38 | (31.1) | 84 | (68.9) |  | 0.632 | 0.236-1.691 | 0.360 | 46 | (37.7) | 76 | (62.3) |  | 1.322 | 0.569-3.070 | 0.517 |
| Non-govt. employee | 165 | (52.4) | 150 | (47.6) |  | 1.545 | 0.686-3.480 | 0.293 | 82 | (26.0) | 233 | (74.0) |  | 0.812 | 0.317-2.082 | 0.664 | 118 | (37.5) | 197 | (62.5) |  | 1.336 | 0.605-2.951 | 0.474 |
| Businessman | 29 | (55.8) | 23 | (44.2) |  | 1.348 | 0.519-3.499 | 0.539 | 19 | (36.5) | 33 | (63.5) |  | 0.496 | 0.170-1.445 | 0.199 | 20 | (38.5) | 32 | (61.5) |  | 1.280 | 0.499-3.285 | 0.608 |
| Unemployedt | 17 | (63.0) | 10 | (37.0) |  | 1 |  |  | 6 | (22.2) | 21 | (77.8) |  | 1 |  |  | 12 | (44.4) | 15 | (55.6) |  | 1 |  |  |
| Family type |  |  |  |  |  |  |  |  |  |  |  |  |  |  |  |  |  |  |  |  |  |  |  |  |
| Nuclear | 809 | (51.5) | 763 | (48.5) | 0.175 | 1.046 | 0.847-1.291 | 0.676 | 612 | (38.9) | 960 | (61.1) | 4.753 | 0.782 | 0.626-0.976 | 0.029 | 707 | (45.0) | 865 | (55.0) | 0.121 | 0.963 | 0.779-1.190 | 0.728 |
| Joint | 234 | (52.6) | 211 | (47.4) | 0.676 | 1 |  |  | 148 | (33.3) | 297 | (66.7) | 0.029 | 1 |  |  | 196 | (44.0) | 249 | (56.0) | 0.728 | 1 |  |  |
| No of family member |  |  |  |  |  |  |  |  |  |  |  |  |  |  |  |  |  |  |  |  |  |  |  |  |
| <5 | 746 | (51.5) | 703 | (48.5) | 0.106 | 1.033 | 0.850-1.254 | 0.745 | 553 | (38.2) | 896 | (61.8) | 0.514 | 0.929 | 0.760-1.136 | 0.473 | 631 | (43.5) | 818 | (56.5) | 3.108 | 1.191 | 0.981-1.447 | 0.078 |
| ≥5 | 297 | (52.3) | 271 | (47.7) | 0.745 | 1 |  |  | 207 | (36.4) | 361 | (63.6) | 0.473 | 1 |  |  | 272 | (47.9) | 296 | (52.1) | 0.078 | 1 |  |  |
| Monthly family income |  |  |  |  |  |  |  |  |  |  |  |  |  |  |  |  |  |  |  |  |  |  |  |  |
| <20,000 | 256 | (50.0) | 256 | (50.0) | 1.412 | 1.066 | 0.861-1.318 | 0.559 | 216 | (42.2) | 296 | (57.8) | 8.219 | 0.732 | 0.589-0.911 | 0.005 | 263 | (51.4) | 249 | (48.6) | 12.147 | 0.707 | 0.571-0.876 | 0.001 |
| 20,000-30,000 | 267 | (53.7) | 230 | (46.3) | 0.494 | 0.918 | 0.740-1.138 | 0.436 | 193 | (38.8) | 304 | (61.2) | 0.016 | 0.842 | 0.674-1.051 | 0.128 | 209 | (42.1) | 288 | (57.9) | 0.002 | 1.029 | 0.828-1.279 | 0.795 |
| >30,000 | 520 | (51.6) | 488 | (48.4) |  | 1 |  |  | 351 | (34.8) | 657 | (65.2) |  | 1 |  |  | 431 | (42.8) | 577 | (57.2) |  | 1 |  |  |
| Residence |  |  |  |  |  |  |  |  |  |  |  |  |  |  |  |  |  |  |  |  |  |  |  |  |
| Rural area | 288 | (47.2) | 322 | (52.8) | 7.083 | 1.295 | 1.070-1.566 | 0.008 | 238 | (39.0) | 372 | (61.0) | 0.665 | 0.922 | 0.758-1.121 | 0.415 | 322 | (52.8) | 288 | (47.2) | 22.733 | 0.629 | 0.520-0.762 | <0.001 |
| Urban area | 755 | (53.7) | 652 | (46.3) | 0.008 | 1 |  |  | 522 | (37.1) | 885 | (62.9) | 0.415 | 1 |  |  | 581 | (41.3) | 826 | (58.7) | <0.001 | 1 |  |  |
| Knowledge |  |  |  |  |  |  |  |  |  |  |  |  |  |  |  |  |  |  |  |  |  |  |  |  |
| More accurate | 0 | (.0) | 974 | (100.0) | — | — | — | — | 372 | (38.2) | 602 | (61.8) | 0.211 | 0.959 | 0.801-1.148 | 0.646 | 424 | (43.5) | 550 | (56.5) | 1.167 | 1.102 | 0.924-1.313 | 0.280 |
| Less accurate | 1043 | (100.0) | 0 | (0.0) | — |  |  |  | 388 | (37.2) | 655 | (62.8) | 0.646 | 1 |  |  | 479 | (45.9) | 564 | (54.1) | 0.280 | 1 |  |  |
| Attitude |  |  |  |  |  |  |  |  |  |  |  |  |  |  |  |  |  |  |  |  |  |  |  |  |
| More positive | 655 | (52.1) | 602 | (47.9) | 0.211 | 0.959 | 0.801-1.148 | 0.646 | 0 | (0.0) | 1257 | (100.0) | — | — | — | — | 521 | (41.4) | 736 | (58.6) | 14.885 | 1.428 | 1.191-1.711 | <0.001 |
| Less Positive | 388 | (51.1) | 372 | (48.9) | 0.646 | 1 |  |  | 760 | (100.0) | 0 | (0.0) | — |  |  |  | 382 | (50.3) | 378 | (49.7) | <0.001 | 1 |  |  |
| Practice |  |  |  |  |  |  |  |  |  |  |  |  |  |  |  |  |  |  |  |  |  |  |  |  |
| More frequent | 564 | (50.6) | 550 | (49.4) | 1.167 | 1.102 | 0.924-1.313 | 0.280 | 378 | (33.9) | 736 | (66.1) | 14.885 | 1.428 | 1.191-1.711 | <0.001 | 0 | (0.0) | 1114 | (100.0) | — | — | — | — |
| Less frequent | 479 | (53.0) | 424 | (47.0) | 0.280 | 1 |  |  | 382 | (42.3) | 521 | (57.7) | <0.001 | 1 |  |  | 903 | (100.0) | 0 | (0.0) | — |  |  |  |

Furthermore, 55.2% of respondents had more frequent practices towards the COVID-19. The rates of more frequent practices were significantly more likely to be (i)female (59.2% vs. 52.6% in male, *p*=.003), (ii) older (age > 30 years) (64.0% vs. 48.6% in aged 12-20 years, *p*<.001), (iii) have higher education (63.6% vs. 35.0% in secondary [6^th^-10^th^ grades], *p*<.001), (iv) be a housewife (68.8% vs. 52.2% in students, *p*=.001), (v) have monthly family income 20,000-30,000 BDT (57.9% vs. 48.6% in those < 20,000, *p*=.002), (vi) be a respondent from urban area (58.7% vs. 47.2% in those from rural areas, *p*<.001), and (vii) have more positive attitudes (58.6% vs. 49.7% in comparatively less positive attitudes, *p*<.001).

The sociodemographic factors of more frequent practices were sex (males vs females: OR=0.76; 95%CI=0.64-0.92, *p*=.003), being younger (12-20 years) vs. older (>30 years)(OR=0.53; 95%CI=0.38-0.75, *p*<.001), having secondary (6^th^-10^th^ grades) vs. higher education(above bachelor) (OR=0.31; 95%CI=0.12-0.79, *p*=.014), having monthly family income less than 20,000 vs. more than 30,000 BDT (OR=0.71; 95%CI=0.57-0.88, *p*=.001), rural vs. urban area (OR=0.63; 95CI%=0.52-0.76, *p*<.001), and having more vs. comparatively less positive attitudes (OR=1.43; 95%CI=1.19-1.71, *p*<.001).

## Discussion

This study was conducted aiming at measuring the level of knowledge, attitude, and practice of COVID-19 and perceptions regarding the disease among Bangladeshi people. The findings reveal a substantial number of sociodemographic factors that affect KAP and should prove useful when planning health education programs about emerging infectious diseases.

In the scope of perception towards COVID-19, the vast majority of the study participants reported some of the commonest symptoms of COVID-19 related [28], with only a very small minority being unaware of any of the symptoms, similar to other studies elsewhere [22,19]. Knowledge about the incubation period was also excellent and similar (86.2%) to the study conducted by Zegarra et al. [22] Similarly, routes of transmission of COVID-19 were reported by the participants: with only a minimal minority (0.2%) participants not being sure or unable of recognizing transmission routes. Perception of COVID-19 severity in the community showed that only 13.8 % did not face any difficulty when they discussed and tried to convince their family members about COVID-19 severity. Most of the responses a by the participants indicated negligence about the severity of the disease, reluctance to use masks, and the reluctance of complying with not being able to stop going out of the house. This may imply less participation in the preventive measures stipulated by the government as well as less inclination to observe social distancing and other individual preventive actions, although some alternative adaptive strategies were also mentioned. The most frequently identified gap in knowledge among participants was related to disease treatment. Only 18.3% of participants believed that there is no treatment for COVID-19, while 47.3% participants indicated that COVID-19 is a treatable disease, similar to another study [29]. Furthermore, only 1%of the participants reported vaccine as an option for preventing COVID-19, in marked contrast with the previous study by Srichan et al which found that 31.2% were aware of the vaccine as a potential option [29]. In an earlier study by Aldowyan et al., only 19% of the participants were aware that there is no treatment for coronavirus like MERS-CoV, while 26.6% indicated the use of supportive treatment for MERS-CoV, and 31.1% of the participants mentioned the vaccine option for preventing MERS-CoV [24].

Compared to 3 others studies [17,29,30], our survey uncovered markedly reduced accurate knowledge, positive attitudes, and frequent practices towards COVID-19 [17,30]. This indicates a significant education gap, likely reflecting suboptimal public health information and dissemination regarding COVID-19, particularly since as indicted our survey primarily sampled educated younger people with ready access to a variety of information sources. Indeed, more accurate knowledge was significantly more likely among young adults, but intriguingly among respondents from rural areas, possibly reflecting that most of the participants were students, and that they all went back home, mostly to rural areas during the lockdown period. Srichanet al. found marital status, education, occupation, annual income were significant factors associated with accurate knowledge of COVID-19 [29], whereas Zhong et al. found that male sex, age-group of 16-29 years, marital status, education, employment and being a student were significantly associated with knowledge [17]. Therefore, tailoring of the information provided by health officials and other media outlets on the disease needs to address the multifactorial nature of the drivers leading to reduced knowledge.

The findings showed virtually universal agreement among the participants towards reporting to health authorities suspected cases of COVID-19, on the issue wearing a face mask before going to a crowded place, and in following other recommendations. These findings were similar to a very recent study conducted in China, during the rapid rise of COVID-19 outbreak [17]. Saqlain et al. also reported positive attitudes among the vast majority of healthcare professionals towards wearing protective gear [29]. Similarly, the overall attitude towards actions such ‘wash hands and face after coming from outside’ and ‘health education can play an important role for COVID-19 prevention’ was universally favorable. Like in this study, Saqlain et al. reported that more than 80% participants strongly agreed that transmission of COVID-19 could be prevented by following universal precautions given by WHO or CDC [30]. During the SARS epidemic, 70.1-88.9% of Chinese residents believed that SARS can be successfully controlled or prevented [17,32]. Zhong et al. found that 90.8% of the respondents agreed that with control measures such as traffic limits all throughout China, and the shutdown of cities and counties of Hubei Province [17]. Surprisingly, the participants’ attitudes differ by age, education, marital status, occupation, family type, monthly income, and practices. In contrast, Saqlain et al. found participants’ attitudes were not affected by age, gender, experience, and job/occupation. Giao et al. also found that attitudes regarding COVID-19 did not present any significant associations with age, gender, and experience, but found a statistically significant association with occupation/job [32]. Also of relevance, Albarrak et al. and Khan et al. did not found any differences in attitude towards MERS among doctors, pharmacists, and nurses [33, 34].

In the multiple logistic regression analyses, sociodemographic variables associated with more positive attitudes regarding COVID-19 were older age, having higher education, being employed, having joint family, having higher monthly family income, and implementing more frequent practices, overall recapitulating previous findings from China [17].

The issue of preventive practices merits some comment since for some measures such as hand washing the results were remarkably similar to the findings others [30,34,35], albeit with the exception of the study by Srichan et al., in which 54.8% did not regularly use soap during washing of hands [29]. Globally, women were significantly more likely to adopt preventive activities than men, a finding that may be of critical importance since targeting of women during household dissemination of education and preventive guidelines may ultimately yield improved implementation in households. Accordingly, we found that the sociodemographic factors associated with more frequent practice measures were being female, older age, having higher education, higher income, urban area residence, and having more positive attitudes. Male gender, occupation of “students”, COVID-19 knowledge score, marital status, and residence were significantly associated factors in the Zhong et al. study, while experience was indicated by Saqlainet al., Hussain et al. and Ivey et al [30,36,37],

Considering the fact that Bangladesh is a multi-ethnic country with vastly different economic income, education levels, traditions, it is expected that the levels of knowledge, attitude, and prevention will also markedly differ in the population. Although good KAP was present in a sizeable proportion of the sample, it is very likely that population sectors that have no access to internet or live in regions with less likely fast escalation of transmission may also display reduced KAP when standard and uniform education and dissemination initiatives are promulgated and implemented. Indeed, it is highly probable that large clusters of people will become less informed and adoptive of prevention practices on COVID-19 [22]. Accessibility to information, dissemination and illustration of preventive behaviors, and sanitary educational measures are essential, especially in rural areas, among old people, poorer neighborhoods or communities, since these may have difficulties in getting access to novel information or encounter financial or resource barriers to implementation of preventive measures [15]. It is common consensus that a more educated population about any given disease will comply better with the preventive and treatment measures [38].

### Limitations

This study has several limitations. First, this study followed a cross-sectional study design. Therefore, causal inferences may not be established. Second, compared with face-to-face interviews, self-reporting has limitations including multiple biases. Third, this study used an online-based survey method to avoid possible transmission, such that the cohort reflects sampling biases by being conducted online, thereby restricted to only those with internet access, and consequently unlikely to represent an accurate reflection of the whole Bangladeshi population. Notwithstanding, our study indicates that KAP assessments towards the COVID-19 pandemic of vulnerable populations warrant special effort to address the gaps incurred by the current study approach. Fourth, we used a limited number of questions to measure the level of knowledge, attitude, and practice. Thus, additional assessments would be important, using all aspects of KAP towards COVID-19, to determine the actual extent of KAP in the general population. Additionally, the unstandardized and inadequate assessment of attitudes and practices towards COVID should be developed via focus group discussion and in-depth interviews and constructed as multi-dimensional measures.

## Conclusion

Our findings indicate that after the immediate lockdown and during the rapid rise period of the COVID-19 outbreak, internet users in Bangladesh displayed substantial differences in KAP regarding the pandemic. Our findings suggest the need for effective and tailored health education programs aimed at improving COVID-19 knowledge, thereby leading to more favorable attitudes and to implementation and maintenance of safe practices.

## Data Availability

All data were included in the manuscript.

https://data.mendeley.com/datasets/s9wgjh7t7f/draft?a=a3c2342c-1eba-45dc-8335-9756f9e14ff4

## Acknowledgments

The authors would like to extend heartfelt graciousness to all the participants who participated in this study voluntarily and spontaneously. Furthermore, the authors highly appreciate the contribution of Rakib Hasan, Lakshmi Rani Kundu, A.S.M MahbubulAlam, Gobida Deb Arya, ArzaMiraz Keya, Md. Abdul Halim, Md Marzan Sarkar, Assaduzzaman Nur, Mohammad Yusuf, Jobair Sami, Md. SanzidMostofa, Miraz Mostafa, Mahmdul Hasan Shoron, Prokriti Biswas, Piya Ferdous, MahirShahariarShowrov, SadmanSakib Samir, Maisha Meherin, SyedaSurayia Sultana, Sayma Islam Alin, RejinaAkter, A H Shourav, KifayatSadmanIshadi, Md. Safiul Hasan, TasnimaAkterTasin, FatemaAkterBethi, Sanzida Amin, Arpita Chakrabarty, SayedaSumaiyaNahrin, RabeyaAkter Mohua, Md. RayhanSakib, Tareq Mahmud, Md. Fakhrul Islam Maruf, Anik Roy, Tariqul Islam, Tasnimul Ahsan Shakhar, and Team SBCC (Social Behavior and Change Communication), during data collection periods.

## Authors contributions

MSI: Conceptualization, Methodology, Investigation, Data curation, Formal analysis, Writing-original draft, Editing, Validation., MZF: Conceptualization, Investigation, Writing-original draft, Editing, Validation. MTS: Editing, Validation., ASMM: Editing, Validation., JAZV: Editing, Validation., DG: Editing, Validation.

